# ICU admissions and in-hospital deaths linked to covid-19 in the Paris region are correlated with previously observed ambient temperature

**DOI:** 10.1101/2020.07.10.20150508

**Authors:** Mehdi Mejdoubi, Xavier Kyndt, Mehdi Djennaoui

## Abstract

**OBJECTIVE:** To study the effect of weather on severity indicators of coronavirus disease 2019 (covid-19).

**DESIGN:** Ecological study.

**SETTING:** Paris region.

**POPULATION:** Severely ill patients with covid-19.

**MAIN OUTCOME MEASURES:** Daily covid-19-related intensive care unit (ICU) admission and in-hospital deaths in the Paris region, and the daily weather characteristics of Paris midtown.

**RESULTS:** Daily ICU admissions and in-hospital deaths were strongly and negatively correlated to ambient temperatures, with a time lag. The highest Pearson correlation coefficients and statistically significant P values were found 8 days before occurrence of ICU admissions and 15 days before deaths.

**CONCLUSIONS:** The study findings show a strong effect of previously observed ambient temperature that has an effect on severity indicators of covid-19.

**Strengths and limitations of this study:** We assessed the link between weather characteristics with a time lag and covid-19-related hospital severity indicators.

This is the first study, with reliable data, to find a strong negative correlation between previously observed ambient temperature with covid-19-related iCU admissions and in-hospital deaths.

It is compatible with the natural history (including hospitalisation) of covid-19 outbreak in a highly developed country.

We also report the duration after which iCU admissions and in-Hospital deaths occurs following temperature variation (respectively median of 8 and 15 days).

Our analyses are mainly valid for temperate climate countries.

## Introduction

The ongoing major epidemic of coronavirus disease 2019 (covid-19) infection is mainly located in cold areas of temperate countries of the northern hemisphere,^1^ which raises the question of weather influence.

Continental France has been severely hit by this outbreak of covid-19, with around 30 000 deaths, including more than 19 000 in-hospital deaths (as of 15 June 2020). The main affected regions being hit were its eastern region (mainly Alsace) and the Paris region (Île de France) with, respectively, more than 3500 and 7200 in-hospital deaths.

Monitoring the outbreak of covid-19 with biological testing is imprecise as the infection can be asymptomatic.^2^ Data such as deaths or intensive care unit (ICU) admissions (reflecting critically ill patients) are more reliable, especially in highly developed countries. In the natural course of infection with covid-19, a pulmonary worsening may appear around 5 to 9 days after infection onset,^2 3^ with an immunological mechanism known as “cytokine storm”^1^ that may lead to a hospitalization. Subsequently, death may occur at different moments (depending on hospitalization, ICU admission, and ventilation). The median duration of hospitalization before death is around 9 days^3^ in covid-19 critically ill patients in a highly developed country, therefore death should occur around 14 to 18 days after the onset of infection, although it can of course happen much later. Weather effect, if existing, could influence a patient’s ICU admission or death the same day of occurrence, and the days before. Once a patient is hospitalized, no weather effect should be seen in highly developed countries if hospital ambient air is regulated (which is the case in France).

To study the hypothesis of a weather effect on severity indicators of covid-19, we tested the correlation between these indicators and weather characteristics in a single area with a high-density population and a heavy burden.

## Methods

### Setting

Paris region (Île de France) has a population of 12 million inhabitants, with the highest population density in France (mainly located in Paris and its suburbs within a 30 km range). The Paris region was chosen as it has the highest death toll in France and is the smallest region (12 012 km^2^). The weather is therefore rather homogeneous throughout the region and can be approximated by the weather in midtown Paris. The healthcare system is characterized by a high density of public and private hospitals with no restriction on access to healthcare. The capacity of ICU beds in France was increased twofold by the end of March 2020, and there was no significant shortage of this kind of beds during the outbreak. Prospectively monitored data (ICU admissions, in-hospital deaths) for covid-19 infection in the Paris region, as everywhere in France, are electronically centralized. A lockdown in France was started on 17 March 2020, and the peak number of deaths was reached on 6 April 2020. A progressive but significant reduction of cases and deaths began after the first week of May. Lockdown was eased on 11 May, with mandatory rules on social distancing.

### Data sources

Daily hospital data (covid-19-related in-hospital deaths, ICU admissions) in the Paris region were retrieved from the national governmental database “Geodes”.^4^ Meteorological factors (hours of sunshine, rainfall, minimal temperature, maximal temperature) were retrieved from the French national meteorology agency.^5^ Mean temperature was estimated using the formula:

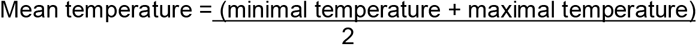

All of these data are public and freely accessible.

### Statistical analysis

Hospital data were correlated to meteorological factors of Paris midtown. A bivariate analysis was performed and Pearson’s correlation coefficients calculated. The significance level was set at 5%. A correlation coefficient >0.7 was considered as very strong, 0.7 to 0.5 as strong, 0.5 to 0.3 as mild, and <0.30 as weak. Meteorological factors were correlated against hospital data on the same day, as death or iCU admission can happen on arrival at the hospital. As weather may have influenced the prognosis of patients before hospitalization, we sequentially tested daily hospital data with the weather observed from 1 to 19 day(s) before ICU admission or death. The period from 31 March 31 to 10 May 2020 was chosen because it corresponds to stabilization of the outbreak following the national lockdown. During this 41-day period, they were 4355 ICU admissions and 5641 deaths in the Paris region (Figure 1). Weather characteristics were therefore observed from 12 March until 10 May. We also assessed a longer period (62 days), with hospital data from 31 March to 31 May and weather data from 12 March to 31 May (a total of 4689 ICU admissions and 6299 deaths).

**Fig 1.**
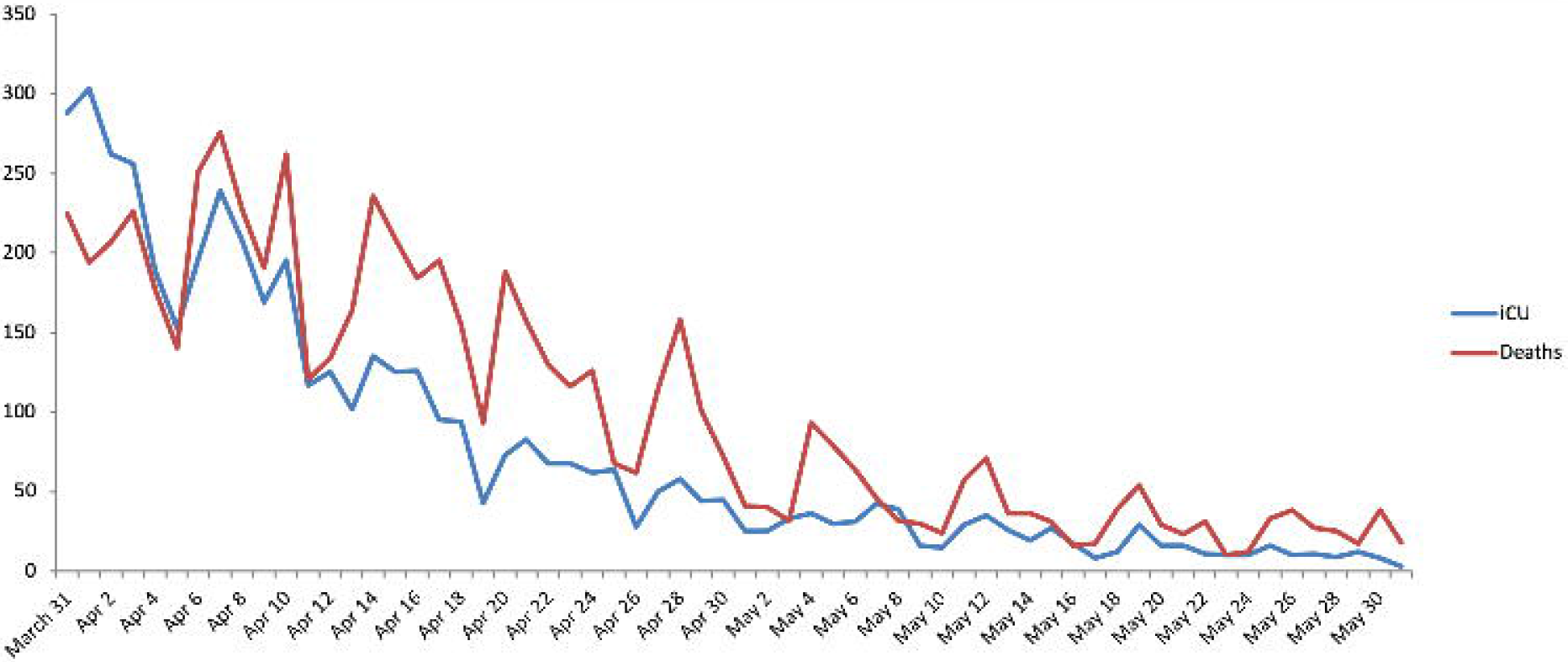
Paris-Region daily covid-19 ICU-Admissions and in-Hospital-Deaths Curves.

The statistical analyses were performed with R software version V4.0.0.

### Patient and public involvement

No patients were directly involved in this study.

## Results

No statistical correlations were found between sunshine or rain and ICU admissions or in-hospital deaths during the 41-day study period. There was a negative correlation with minimal, mean, and maximal temperatures with both outcomes (Figure 2). This correlation was considered strong/very strong (with all three temperatures) for ICU admissions from day –12 to day –6, with the highest correlation at day –8. This correlation was considered strong/very strong for in-hospital deaths from day −16 to day −11 (with all three temperatures) with the highest correlation at day −15. Correlations were statistically significant for most of the considered period Table 1), even more so for correlations between ICU admissions and temperature (Table 2).

**Table 1.**
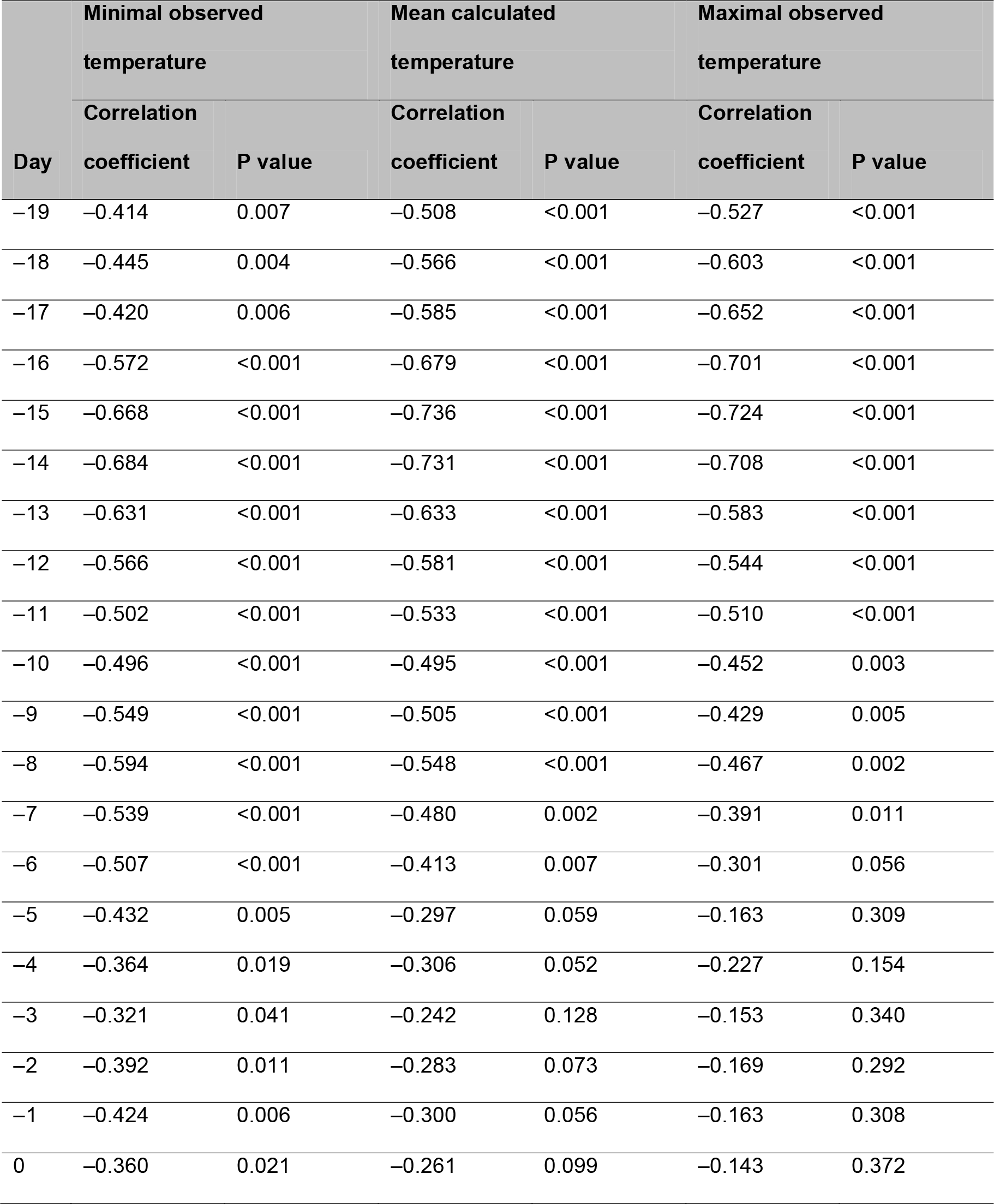
Correlations between in-hospital covid-19-related deaths in the Paris region and temperatures measured from 19 days before death to the date of death (study period: 31 March 31 to 10 May 2020)

**Table 2.** Correlations between covid-19-related ICU admissions in the Paris region and temperatures measured from 19 days before ICU admission to the date of admission (study period: 31 March to 10 May 2020)

| Day | Minimal observed temperature |  | Mean calculated temperature |  | Maximal observed temperature |  |
| --- | --- | --- | --- | --- | --- | --- |
|  | Correlation |  | Correlation |  | Correlation |  |
|  | coefficient | P value | coefficient | P value | coefficient | P value |
| -19 | -0.246 | 0.121 | -0.430 | 0.005 | -0.523 | <0.001 |
| -18 | -0.311 | 0.048 | -0.493 | 0.001 | -0.582 | <0.001 |
| -17 | -0.333 | 0.034 | -0.520 | <0.001 | -0.609 | <0.001 |
| -16 | -0.409 | 0.008 | -0.554 | <0.001 | -0.615 | <0.001 |
| -15 | -0.451 | 0.003 | -0.57 | <0.001 | -0.610 | <0.001 |
| -14 | -0.449 | 0.003 | -0.552 | <0.001 | -0.586 | <0.001 |
| -13 | -0.471 | 0.002 | -0.561 | <0.001 | -0.583 | <0.001 |
| -12 | -0.528 | <0.001 | -0.631 | <0.001 | -0.656 | <0.001 |
| -11 | -0.609 | <0.001 | -0.699 | <0.001 | -0.705 | <0.001 |
| -10 | -0.685 | <0.001 | -0.733 | <0.001 | -0.706 | <0.001 |
| -9 | -0.752 | <0.001 | -0.749 | <0.001 | -0.685 | <0.001 |
| -8 | -0.789 | <0.001 | -0.767 | <0.001 | -0.687 | <0.001 |
| -7 | -0.755 | <0.001 | -0.709 | <0.001 | -0.611 | <0.001 |
| -6 | -0.703 | <0.001 | -0.657 | <0.001 | -0.561 | <0.001 |
| -5 | -0.632 | <0.001 | -0.581 | <0.001 | -0.482 | 0.001 |
| -4 | -0.604 | <0.001 | -0.568 | <0.001 | -0.478 | 0.002 |
| -3 | -0.590 | <0.001 | -0.546 | <0.001 | -0.449 | 0.003 |
| -2 | -0.655 | <0.001 | -0.589 | <0.001 | -0.472 | 0.002 |
| -1 | -0.641 | <0.001 | -0.557 | <0.001 | -0.420 | 0.006 |
| 0 | -0.569 | <0.001 | -0.465 | 0.002 | -0.313 | 0.046 |

ICU, intensive care unit.

**Fig 2.**
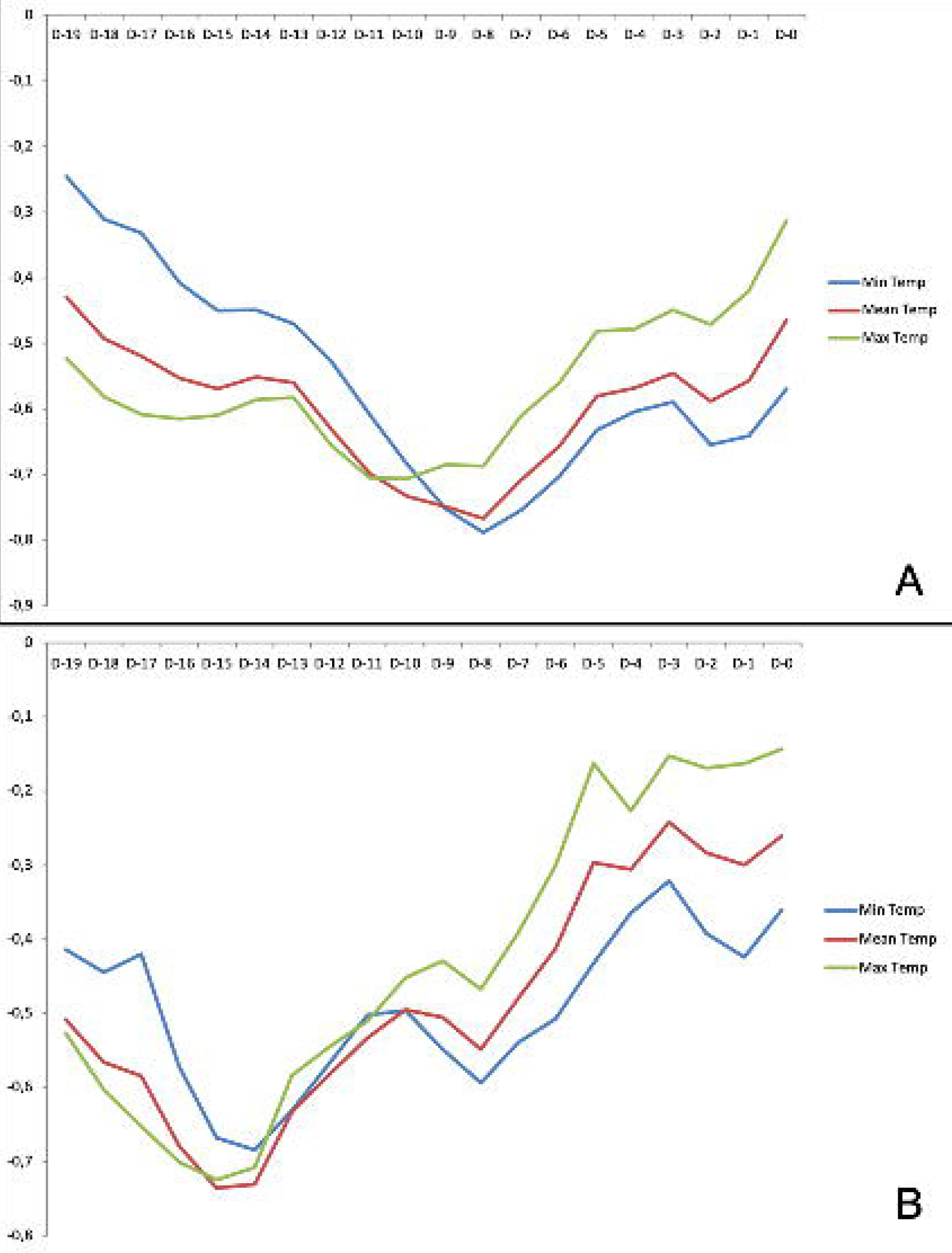
Correlation coefficients of midtown Paris temperatures and (A) daily ICU admissions and (B) in-hospital deaths in the Paris region. In ordinate is Correlation coefficient (0 to –1), in abscissa (days D –19 to 0).

A temperature drop preceded the peak of ICU admissions and in-hospital deaths (Figure 3), with the highest correlation coefficients (−0.767 and −0.736, respectively, for mean temperature) corresponding to a time lag of 8 and 15 days, respectively. There was a second, smaller peak for ICU admissions at day –2, and a second peak of correlated in-hospital deaths at day –8.

**Fig 3.**
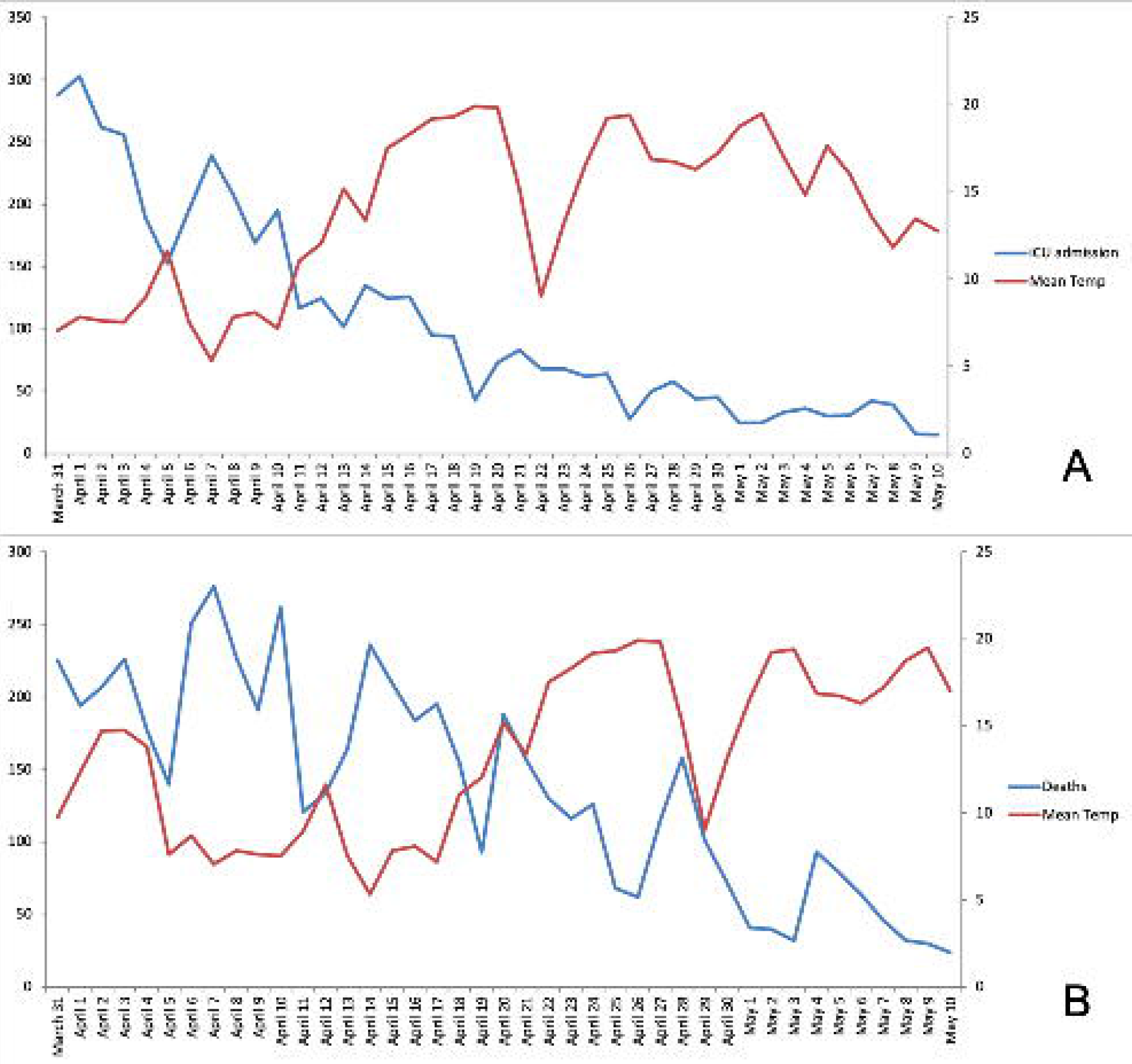
Evolution of (A) ICU admissions and (B) daily death tolls superimposed with mean environmental temperatures observed 8 days before ICU admission and 15 days before death (as these showed the highest correlation coefficients). Left ordinate is daily toll (0–350), right ordinate is temperature (0–25 °C).

The longer study period (62 days) also showed a negative correlation between severity indicators and all three temperatures (maximal correlation coefficients at around –14 to –15 days for deaths and –8 days for ICU admissions) with significant P values, but with weaker correlation coefficients (data not shown).

## Discussion

The findings of this study suggest that severity indicators of the covid-19 outbreak strongly correlated with ambient temperatures in the Paris region. France has a mainly oceanic climate, which is shared by most of Western Europe (UK, Belgium, Netherlands, and Germany).

Many early studies,^6-9^ from different continents and settings, have linked severe acute respiratory syndrome coronavirus 2 (SARS-Cov-2) transmission to meteorological factors. Each 1 °C increase in average ambient temperature and in diurnal temperature range reduced the number of daily cases in China,^6^ whereas a specific absolute humidity of around 6 g/m^3^ favoured transmission. Prata et al.^7^ described a significant linear negative relationship between confirmed cases of covid-19 and ambient temperature from 16.8 °C to 25.8 °C (with a threshold above 25.8 °C where the infection curve became flat). Also, the epidemic intensity decreased following days at a higher temperature (with a time lag), and the peak of infections was at approximately 10 °C.^8^ This finding does not necessarily suggest that lower temperature *per se* affects the virus, as it also modifies human activities. The interactions between ambient temperature, humidity, wind, and hours of sunshine are complex, and are not the purpose of this article. Sajadi et al.^9^ found that relative humidity (44% to 84%) and mean temperature (5 °C to 11 °C) before the first deaths were similar in February in eight different cities (which later became the main hotspots of the outbreak at that time). Most of these studies, however, relied on biologically confirmed infections, which are known to depend widely on the biological testing policy and the ability to do it. In France, as elsewhere, biological testing was very limited at the beginning of the pandemic, and only approximately 200 000 patients were confirmed positive, whereas between 3 and 6 million patients were likely to have been infected.

Ma et al.,^10^ in a study from Wuhan, reported a significant negative association with ambient temperature and absolute humidity on daily deaths from covid-19. However, although they tested a lagged effect, the maximum lag was only 5 days between the environmental factors tested and the day of death. This does not fit with the course of pathology from this disease, as death often occurs later (and much later in case of hospitalization, which extends patient survival, at least in highly developed countries). They did not find any effect of air pollutants on death, but some authors hypothesized that pollution could favour spread of the virus.^11^ Death monitoring may also be imprecise, but is quite reliable in highly developed countries and we sought to work on reliable data from France. With regards to the death toll, we chose to focus on in-hospital deaths, which are more precise for the date of occurrence and the cause. Hospital admission thresholds differ among countries, and depend on hospital beds per capita, ventilator availability, and access to healthcare. Within a country, it depends on patient categorization (mainly age and disease severity). Consistent with this, we found different day peaks of death, consistent with the course of hospitalization and the different types of patients. Between 34.6% and 55.9% of patients with covid-19 are hospitalized, but are not admitted to the ICU or have to undergo mechanical ventilation.^12^ The smaller peak observed at day –8 for deaths likely corresponds to fragile hospitalized patients who die early. Some patients are not even admitted to the ICU for ventilation therapy, which is often a single “yes or no” decision. We found that ambient temperatures at day –8 correlated most strongly with ICU admissions, whereas temperatures at day –15 correlated most closely with in-hospital deaths. The difference of 7 days therefore corresponds to the median time between ICU admission and in-hospital death in Paris region.

Humidity and temperature influence survival of influenza virus, SARS-CoV, and MERS-CoV, as well as droplet stabilization, propagation in nasal mucosa, human immunity,^9 13^ and human way-of-life (i.e. indoor and outdoor activities). Therefore, a respiratory outbreak has multifactorial drivers and the number and importance of each of these factors is not only unknown, but may also vary between countries and climes. Models show that while climate may influence this outbreak and/or its severity in a particular location, other factors such as population immunity^13^ are crucial for a potential seasonality. Also, public health controls (school closures, restricting mass gatherings, social distancing) play a major role and likely have a greater effect compared with climate.^14^

Although our results clearly indicate a link between temperature and severity indicators of covid-19, our study does not provide clear answers to the precise effect of temperature. It may be responsible for a greater number of infections that eventually lead to a greater number of deaths, or it may only be responsible for more severe infections, or it may affect both.

### Strengths and limitations

Our study has several strengths. We were able to study reliable data as in-hospital deaths are electronically centralized in France. Lockdown was uniformly applied in France, and was respected by the population, allowing a period of stabilization in April and May. Our study also has some limitations. First, we chose not to study the effect of humidity, which varies more than temperature (between 40% and 90%), depending on the location within the Paris region. Second, the study period is limited to 2 months, but the natural course of this outbreak in France does not allow a longer period as it lasted for only 3 months and the number of infections is currently low. Finally, our conclusions are valid mainly for temperate countries of the northern hemisphere with a highly developed and universal health system.

## Conclusion

Across Europe and the rest of the world, several factors potentially contribute to the severity of this pandemic. Our findings suggest that weather may contribute to the rise and fall of this outbreak in temperate countries of the northern hemisphere and this could indicate a likely resurgence next winter. Therefore, social distancing should be maintained on a long-term basis. Temperature is a strong driver of this outbreak and may influence both viral spread and case fatality. Our results also indicate that covid-19 deaths may increase soon in the southern hemisphere, with an austral winter.

## Data Availability

all data are publicly available:
https://geodes.santepubliquefrance.fr/#c=indicator&i=covid_hospit_incid.incid_dc&s=2020-05-22&t=a01&view=map2.
http://www.meteofrance.com/climat/meteo-date-passee.

https://geodes.santepubliquefrance.fr/#c=indicator&i=covid_hospit_incid.incid_dc&s=2020-05-22&t=a01&view=map2.

## Credit authorship contribution statement

Mehdi Mejdoubi: Conceptualization, Methodology, Software, Data curation, Writing -- original draft. Xavier Kyndt: Methodology, review editing. Mehdi Djennaoui: Methodology, Software, Data curation, review editing.

